# Baseline and Change in Peripheral Blood Transcriptome Enhance Outcome Prediction in Fibrotic Hypersensitivity Pneumonitis: A Prospective Multicenter Cohort Study

**DOI:** 10.1101/2025.11.13.25340198

**Authors:** Evans R. Fernández Pérez, Sonia M. Leach, Stephen M. Humphries, David A. Lynch, Teng Moua, Sachin Chaudhary, Traci N. Adams, Ayodeji Adegunsoye, Mary Beth Scholand, Namita Sood, Brian Vestal

## Abstract

**Rationale:** Data on the prognostic value of peripheral blood mononuclear cell (PBMC) expression profiles, when used in fibrotic hypersensitivity pneumonitis (fHP) patients as an adjunct to traditional clinical assessment in predicting disease progression, remains limited.

**Objectives:** To determine whether a baseline and time-course transcriptomic signature in PBMC from patients with fHP can enhance progression-free survival (PFS) prediction and complement clinical risk stratification.

**Methods:** The prospective multicenter study cohort included 133 participants with fHP. Lasso regression was employed to create a baseline and time-course (baseline to 12-month change) gene signature. We developed multivariable models incorporating clinical variables (age, sex, smoking status, exposure history, FVC% and quantitative measurements of lung fibrosis—derived from data-driven textural analysis, DTA) both with and without the baseline and time-course gene expression, and evaluated these models using receiver operating characteristic curves. Kaplan-Meier curves displaying high and low gene expression risk scores are presented.

**Results:** The addition of a baseline gene expression profile to the logistic regression model of 24-month PFS using baseline clinical parameters markedly improved the predictive accuracy, increasing the area under the curve (AUC) from 0.77 to 0.93. Similarly, a logistic regression model of 12-month PFS using age, sex, smoking status, exposure history, and changes in FVC% and DTA over 12 months had an AUC of 0.81, which improved to 0.95 with the inclusion of changes in gene expression over the same period. A difference in PFS was noted when the cohort was divided based on whether their gene principal component analysis (PCA) scores were above or below the median. At baseline, patients with higher gene PCA scores had lower median survival than those with lower scores (267 days vs. not available; *P* = 0.008). Similarly, when assessing the change in gene expression from baseline to 12 months, patients with higher gene PCA scores had lower median survival than those with lower scores (203 days vs. 449 days; *P* = <0.001).

**Conclusions:** In fHP, a risk-indicative gene expression signature in peripheral blood at baseline, along with its changes over 12 months, predicts disease progression at 24 months and during the subsequent 12 months, respectively. Prognostic models based on gene expression and clinical factors strongly outperform models based solely on clinical factors.

## BACKGROUND

Among patients with hypersensitivity pneumonitis (HP), a complex and immunologically mediated lung disease caused by repeated inhalational exposure to various antigens, early detection of disease progression is crucial, especially in those with pulmonary fibrosis (fHP). Identifying these patients before pulmonary fibrosis progresses—the leading cause of morbidity and mortality—can facilitate personalize counseling and inform treatment strategies.

While progress has been made in understanding the prognostic value at time of diagnosis of clinical,^1, 2^ radiological,^1, 3, 4^ and pathological manifestations ^5, 6^ of progressive fHP none of these clinical measures account for the dynamic biological state that prognostically distinguishes individuals with HP.

The transcriptional profile can serve as a proxy for the biological state, thereby facilitating a deeper understanding of complex diseases like HP. Given the comprehensive nature of the transcriptional profile, it is possible to identify patterns of expression that reflect subtle distinctions in biology that ultimately predict disease outcome and potentially enhanced the prognostic accuracy beyond that of clinical variables alone.

Due to the critical need for prognostic risk stratification in understanding the disease course, we profiled the blood transcriptome in 37 patients with progressive and non-progressive HP with and without lung fibrosis according to change from initial presentation in functional and radiological data within 24 months and survival.^7^ We found that distinct clinical clusters of patients can be separated into transcriptomic subgroups that correlate with survival, and that classification models based on gene expression alone or in addition to clinical factors strongly outperform models based solely on clinical factors, demonstrating the value of molecular phenotyping to bolstering outcome prediction and complementing traditional clinical risk stratification for HP.

In this study, we hypothesize that risk-indicative gene expression signatures characterize the fHP clinical course and can be used to refine prognosis and predict clinical outcomes in this complex disease. Therefore, we evaluated the accuracy of a baseline and a time-course transcriptomic signature, both individually and in combination with traditional clinical measures, in predicting disease progression in a multicenter cohort of patients with fHP.

## METHODS

### Study design and participants

The study cohort consisted of a multicenter 24-month prospective observational study conducted from December 2020 to June 2025 across seven centers in the U.S. involving 137 patients diagnosed with fHP (NCT04844359),^8^ The study was approved by the Mayo Clinic Central Institutional Review Board (#20-004479) and all relying local Institutional Review Boards before enrollment at each site. All patients provided written informed consent before participating in the study.

### Assessments

For inclusion, adult patients were required to have a multidisciplinary diagnosis of fHP according to diagnostic guidelines. They received standard care for fHP, as determined by their treating physicians. Demographics, smoking history, occupational and environmental history, pre-bronchodilator pulmonary function testing, chest HRCT scan, and treatment data were collected at the time of blood draw during study enrollment and at 12 months.

Noncontrast chest HRCT scans, obtained within 60 days of the baseline and at 12 months, were selected for analysis. All HRCT scans were independently reviewed by an experienced thoracic radiologist (D.A.L.) who was blinded to each patient’s clinical status, demographic data, and HRCT date. All scans were classified according to three morphologic categories as outlined by the HP guidelines: typical, compatible or indeterminate for fHP.^9^ We utilized data-driven texture analysis (DTA) to quantify the extent of lung fibrosis on HRCT, shown to be an accurate and reproducible measure of disease severity and progression.^10–13^

### Next Generation Sequencing of Whole Transcriptome RNA-seq

Peripheral blood was collected from consenting patients at each center during study visits. Total RNA was isolated from peripheral blood mononuclear cells (PBMCs) and processed at NJH for next-generation sequencing (NGS) library construction, using the Illumina HiSeq 2500 NGS platform (San Diego, CA, USA) for analysis. Quality control and quantification of the library were performed with an Agilent Technologies Bioanalyzer (Santa Clara, CA, USA) and a Qubit instrument (Thermo Fisher Scientific, Waltham, MA, USA).

### Differential Expression (DE) Analysis

FASTQ files from the Illumina bcl2fastq v2.17 converter were adapter trimmed using skewer v0.2.2 and aligned to the hg19 human reference genome with the STAR aligner v2.4.1^14^ using Ensembl v75 (http://ensembl.org) gene models. Counts of uniquely mapped reads per gene were quantified using the Subread v1.5.1 software. Subject data were scaled using the variance stabilizing transform in the DESeq2 package and visualized using the pheatmap package and principal components analysis (PCA) princomp package (http://www.R-project.org/). Differential gene expression between progressors and non-progressors at baseline was performed with the limma R package^15^ for transcripts with ≥5 counts per million in at least 50% of the samples, using the moderated t-tests and a false discovery rate adjustment to account for multiple comparisons. A second model was also fit but instead of comparing baseline expression values, we compared changes from baseline to 12 months between progressors and non-progressors, again using limma. Results from both models are presented via volcano plots. Pathway analyses were done using Gene Ontology (GO) terms via the enrichR R package.^16^

### Predictive Modeling

Disease progression was defined as the time from blood draw to a decline in FVC or DLCO of ≥10%, respiratory related hospitalization, lung transplantation or death. Individuals were classified as progressors or non-progressors if they had any of these events occur within the first two years of follow up time. To explore the power of gene expression to predict progression status, we utilized a logistic regression approach. First, we fit models using the Least Absolute Shrinkage and Selection Operator (LASSO) regularization where all genes that passed the CPM filter described above were included simultaneously using the glmnet R package.^17^ An optimal penalty parameter that minimized the 10-fold cross validation misclassification rate was identified, and then a final model was fit using this value. The expression values for the genes with non-zero regression coefficients in this model were then isolated, and then the first principal component of the scaled and centered expression was calculated for each sample as a univariate expression score.

To compare the predictive accuracy of the gene score to standard clinical variables, two more logistic regression models were fit. In the reduced model, the covariates included were age, sex, known antigen exposure (yes/no), smoking status (former/never), FVC% at baseline, and DTA fibrosis score at baseline. A second model was fit using all of these same covariates plus the gene expression PCA score. Comparisons were done using the p-values for the regression coefficients as well as Receiver Operator Curves (ROCs) and Area Under the ROC (AUC) calculated using the ROCR R package.^18^ Sensitivity and specificity were also calculated using a predicted probability of being a progressor over 50% from the logistic regression models as the threshold.

This whole procedure was then repeated using change in gene expression from baseline to 12 months in place of baseline gene expression. That is, LASSO logistic regression was used to identify a set of genes where change in gene expression was predictive of progression in the following time period. The first principal component of the change in these genes was used as an expression score in the subsequent models. Two other logistic regression models were fit to evaluate any improvement over using 12-month change in clinical covariates in the same fashion as above with p-values and AUC presented for the two models.

Kaplan-Meier curves with a high and low gene expression score designation, which corresponds to above or below the median value in this cohort are presented.

## RESULTS

Of the 137 eligible patients with fHP, 133 were included in the baseline analyses and the analysis of the 12-month change in gene expression. The study cohort had a mean age of 68.4 years (± 9.4), with an equal sex distribution. 42.1% were ever smoker and 57.1% had an identifiable inciting antigen exposure. The mean FVC was 75.3% (± 19.5), and the mean DLCO was 64.5% (± 21.5). CT appearances were characterized as typical pattern in 24.8% of cases, a compatible pattern in 40.6%, and an indeterminate pattern in 34.5%. The mean baseline DTA fibrosis score was 23.1 (± 17.9). 70 patients had progression-free survival event during the study period (**Table 1**).

**Table 1.** Baseline Characteristics of Primary and Validation Cohorts.

| Characteristic | Study Cohort<br>(N=133) |
| --- | --- |
| Age, years, mean (SD) | 68.4 (9.4) |
| Female, n (%) | 67 (50.3) |
| Ever smoker, n (%) | 56 (42.1) |
| Identifiable antigen exposure, n (%) | 76 (57.1) |
| Pulmonary function, mean (SD) |  |
| FVC% predicted | 75.3 (19.5) |
| DLCO% predicted | 64.5 (21.5) |
| HRCT HP pattern confidence, n (%) |  |
| Typical | 33 (24.8) |
| Compatible | 54 (40.6) |
| Indeterminate | 46 (34.5) |
| DTA total fibrosis extent, mean (SD) | 23.1 (17.9) |
| Disease progression, n (%) | 70 (52.6) |

Among 10,846 genes analyzed, volcano plots using a threshold of an unadjusted p-value of 0.005 revealed 119 genes that met this criterion at baseline and 26 genes for the 12-month change in differential expression between progressors and non-progressors (**Figures 1a** and **1b**).

**Figure 1.**
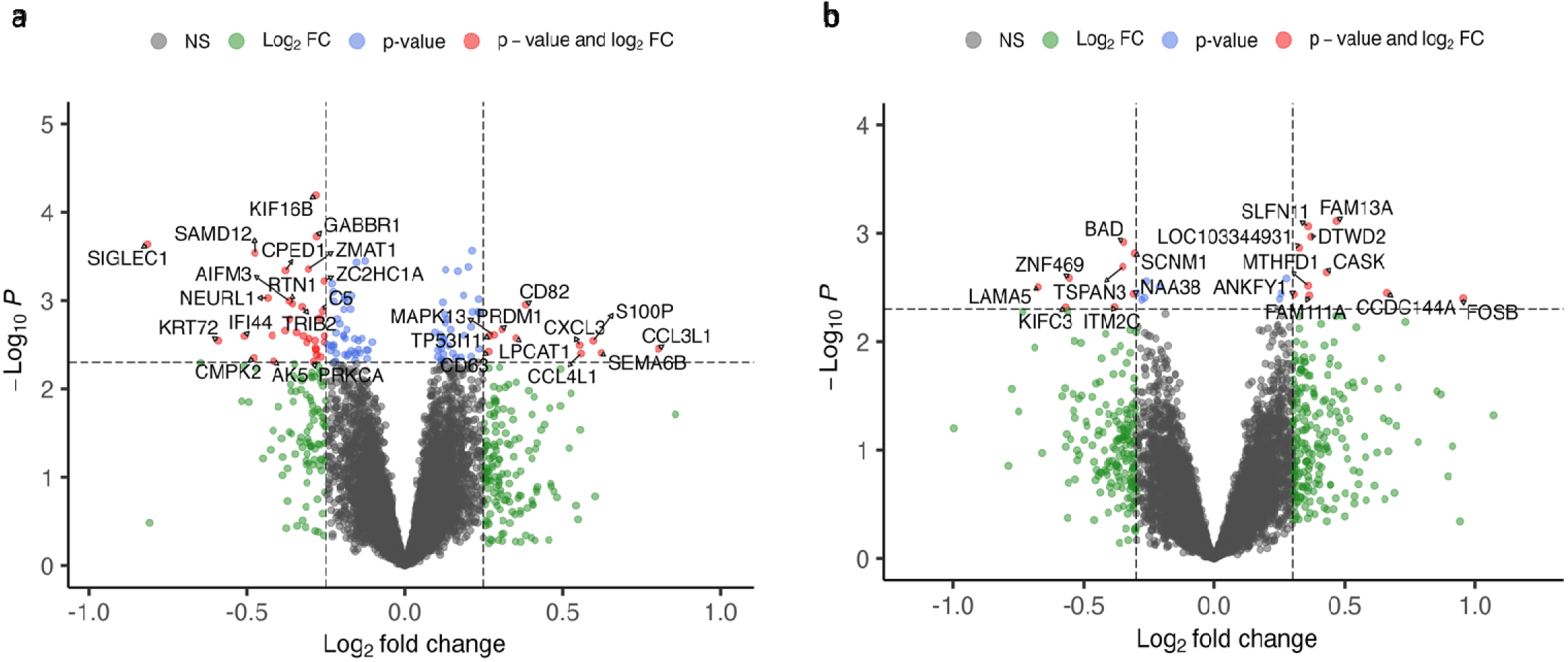
Volcano plot of genes between progressors and non-progressors. Panel a has the differences in mean expression at baseline, and panel b has the differences in the 12-month changes. The dashed horizontal line corresponds to a raw p-value of 0.005, while the vertical line is at a log fold change of 0.3. Downregulated genes on the left and upregulated genes on the right. FDR=false discovery rate.

The biological process terms of GO of the top 300, based on p-value, up- and down-regulated genes in progressors compared to non-progressors at baseline are shown in **Figures 2a** and **2b**. GO terms associated with genes at baseline were mainly related to upregulation of protein phosphorylation, protein modification process, regulation of cell motility and macroautophagy processes, and predominant downregulation of interleukin 27 mediated signaling pathways. Pathways associated with the top 300 genes that had higher change from baseline to 12 months between progressors and non-progressors (**Figure 2c**) included upregulation of cytosolic and vacuolar transport cellular processes.

**Figure 2.**
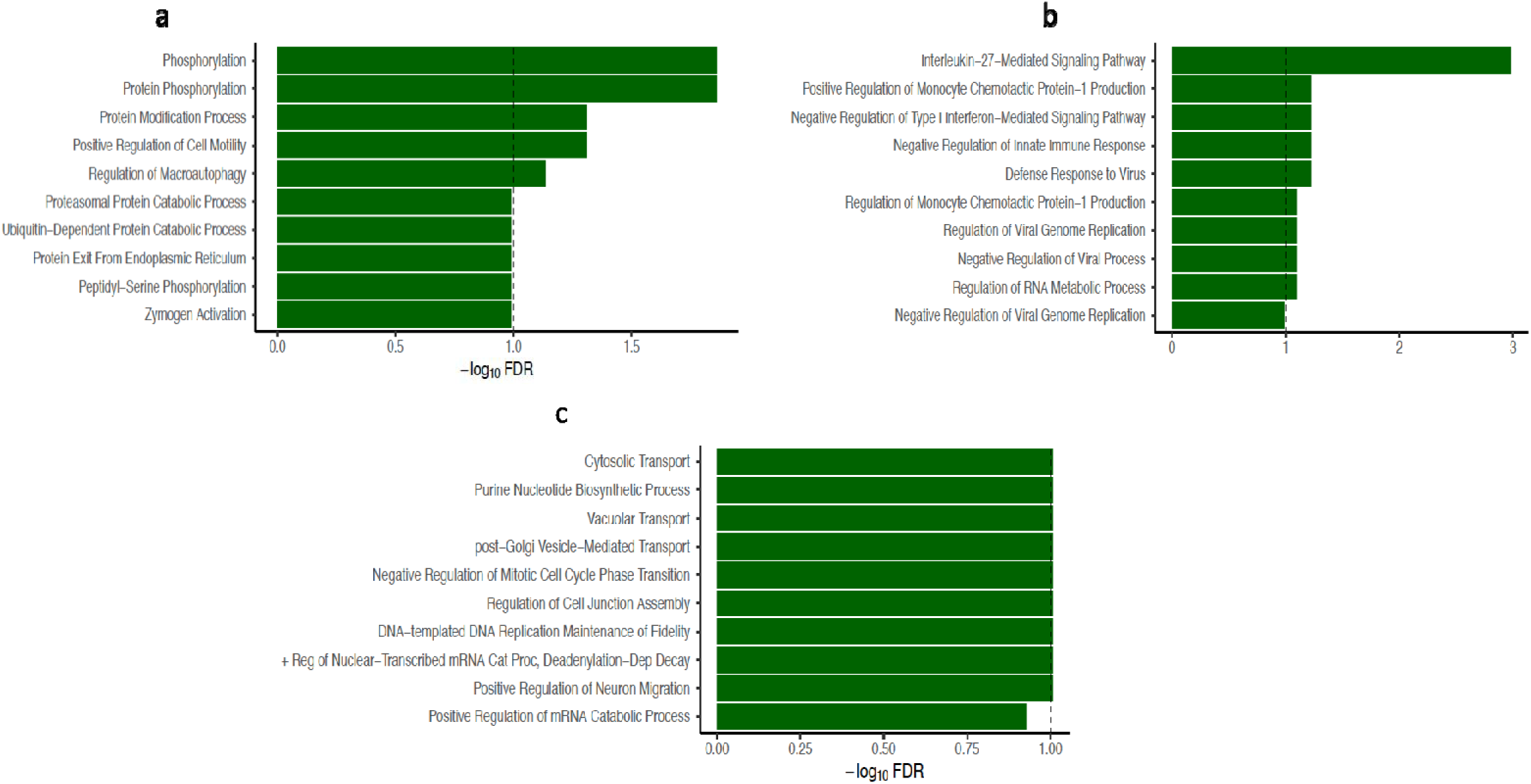
Functional enrichment analysis of the gene ontology pathways of the top 300 up (**a**) and down (**b**) differentially expressed genes at baseline between the progressors and non-progressors. (**c**) Gene ontology pathways associated with change from baseline to 12 months in differentially expressed genes between progressors and non-progressors.

20 genes were selected by the LASSO logistic regression model for the baseline expression signature differentiating progressors from non-progressors: AIFM3, ANKMY2, CCDC61, CCL3L1, CPED1, EML3, GSTM1, HADH, KIF16B, KRT72, PRDM1, RFC3, SAMD12, SIGLEC1, SURF4, TIMM10, UTP14A, VGLL4, ZC2HC1A and ZNF300. Three were selected by LASSO for the change in genes expression from baseline to 12 months: DTWD2, FAM13A and SLFN11. DTA (odds ratio = 1.048, 95% CI 1.015 to 1.083, *P* = 0.004) was the strongest baseline clinical predictor of 24-month progression (**Table 2**). However, no clinical variables were individually significantly predictive of subsequent 12-month progression (**Table 3**).

**Table 2.** Multivariate logistic regression models of baseline clinical variables and gene principal component analysis score.

|  | OR | 95% CI | P value |
| --- | --- | --- | --- |
| **Baseline clinical variables** |  |  |  |
| Age, years | 1.026 | (0.976, 1.079) | 0.317 |
| Sex, male | 0.784 | (0.313, 1.962) | 0.603 |
| Smoking status | 2.383 | (0.954, 5.957) | 0.063 |
| Known antigen exposure | 0.609 | (0.245, 1.512) | 0.285 |
| FVC% | 0.984 | (0.957, 1.011) | 0.237 |
| DTA fibrosis score | 1.045 | (1.012, 1.079) | 0.008 |
| **Baseline clinical variables and gene PCA score** |  |  |  |
| Age, years | 0.957 | (0.889, 1.030) | 0.239 |
| Sex, male | 0.422 | (0.109, 1.633) | 0.212 |
| Smoking status | 2.847 | (0.797, 10.168) | 0.107 |
| Known antigen exposure | 0.303 | (0.075, 1.230) | 0.095 |
| FVC% | 0.993 | (0.956, 1.032) | 0.72 |
| DTA fibrosis score | 1.061 | (1.011, 1.112) | 0.015 |
| Gene PCA score | 4.633 | (2.484, 8.639) | $1.40 \times 10^{-06}$ |

FVC = forced vital capacity; DTA = data-driven textural analysis; PCA = principal component analysis.

**Table 3.** Multivariate logistic regression models of baseline and 12 months change in clinical variables and gene principal component analysis score of change in gene expression from baseline to 12 months.

|  | OR | 95% CI | P value |
| --- | --- | --- | --- |
| **Baseline and 12 months change in clinical variables** |  |  |  |
| Age, years | 1.13 | (1.000, 1.277) | 0.05 |
| Sex, Male | 0.305 | (0.052, 1.776) | 0.186 |
| Known antigen exposure | 2.835 | (0.465, 17.272) | 0.258 |
| Smoking status | 3.197 | (0.649, 15.758) | 0.153 |
| Delta FVC% | 1.036 | (0.911, 1.177) | 0.59 |
| Delta DTA | 1.219 | (0.978, 1.519) | 0.078 |
| **Baseline and 12 months change in clinical variables and gene PCA score** |  |  |  |
| Age, years | 1.178 | (0.918, 1.512) | 0.198 |
| Sex, Male | 0.483 | (0.040, 5.827) | 0.567 |
| Known antigen exposure | 3.021 | (0.280, 32.571) | 0.362 |
| Smoking status | 23.91 | (0.976, 585.897) | 0.052 |
| Delta FVC% | 1.172 | (0.916, 1.499) | 0.206 |
| Delta DTA | 1.387 | (0.985, 1.951) | 0.061 |
| Gene PCA score | 38.732 | (1.647, 910.886) | 0.023 |

FVC = forced vital capacity; DTA = data-driven textural analysis; PCA = principal component analysis.

The AUC for the logistic regression model of progression using baseline clinical parameters of age, sex, smoking status, exposure history, FVC% and DTA was 0.77 (**Figure 3a)**. This was significantly improved when adding the first PC of the baseline expression data for the genes identified in the LASSO model as the AUC increased to 0.93 in combination with baseline clinical variables (**Figure 3b)**. When classifying progression status using a 50% or greater predicted probability from the logistic regression models, the sensitivity, specificity, positive predictive value (PPV) and negative predictive value (NPV) for the clinical variables only were 0.81, 0.67, 0.76, and 0.73 respectively. Again, all values were substantially improved with the addition of gene expression information as sensitivity increased to 0.90, specificity was 0.77, PPV was 0.84, and NPV was 0.86. Similarly, the logistic regression model of progression using age, exposure history, and change in FVC% and change in DTA over 12 months had an AUC of 0.81 (**Figure 3c**), but that was improved to 0.95 (**Figure 3d**) with the addition of change in expression for the genes selected by the LASSO model. The performance characteristics of the model also improved (**Table 4**).

**Figure 3.**
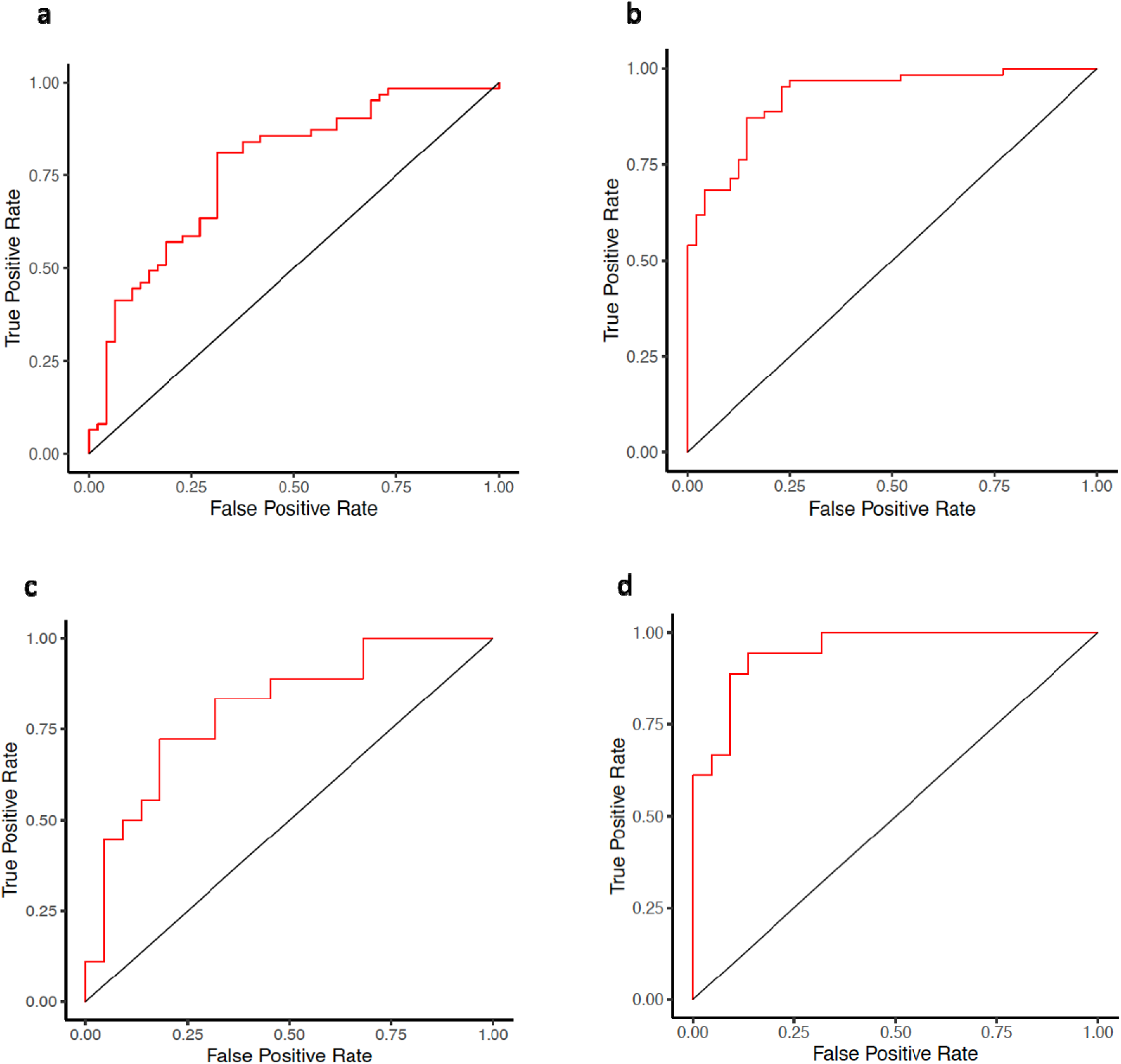
Receiver operating characteristic curves for (**a**) baseline clinical variables (age, sex, exposure history, smoking status, FVC%, and DTA, AUC 0.77), (**b**) baseline clinical variables and genes (AUC 0.93), (**c**) clinical variables (baseline age, sex, exposure history, smoking status, and change from baseline to 12 months in FVC% and DTA, AUC 0.81), (**d**) clinical variables (baseline age, sex, exposure history, smoking status, and change from baseline to 12 months in FVC% and DTA) and change in gene expression from baseline to 12 months (AUC 0.95). AUC = area under the curve; FVC = forced vital capacity; DTA = data-driven textural analysis; PCA = principal component analysis.

**Table 4.** Model performance characteristics.

|  | **Baseline clinical variables** | **Baseline clinical variables and gene PCA score** |
| --- | --- | --- |
| Sensitivity | 0.809 | 0.904 |
| Specificity | 0.666 | 0.770 |
| PPV | 0.761 | 0.838 |
| NPV | 0.727 | 0.860 |
|  | **Baseline and 12 months change in clinical variables** | **Baseline and 12 months change in clinical variables and gene PCA score** |
| Sensitivity | 0.555 | 0.833 |
| Specificity | 0.818 | 0.909 |
| PPV | 0.714 | 0.882 |
| NPV | 0.692 | 0.869 |

NPV = negative predictive value; PCA = principal component analysis score; PPV = positive predictive value.

Differential progression-free survival was observed when the cohort was stratified by gene PCA scores above or below the median in this cohort. At baseline, patients with higher gene PCA scores had lower median survival than those with lower scores (267 days vs. not available; *P* = 0.008, **Figure 4a**). Similarly, when assessing the change in gene expression from baseline to 12 months, patients with higher gene PCA scores had lower median survival than those with lower scores (203 days vs. 449 days; *P* = <0.001, **Figure 4b**).

**Figure 4.**
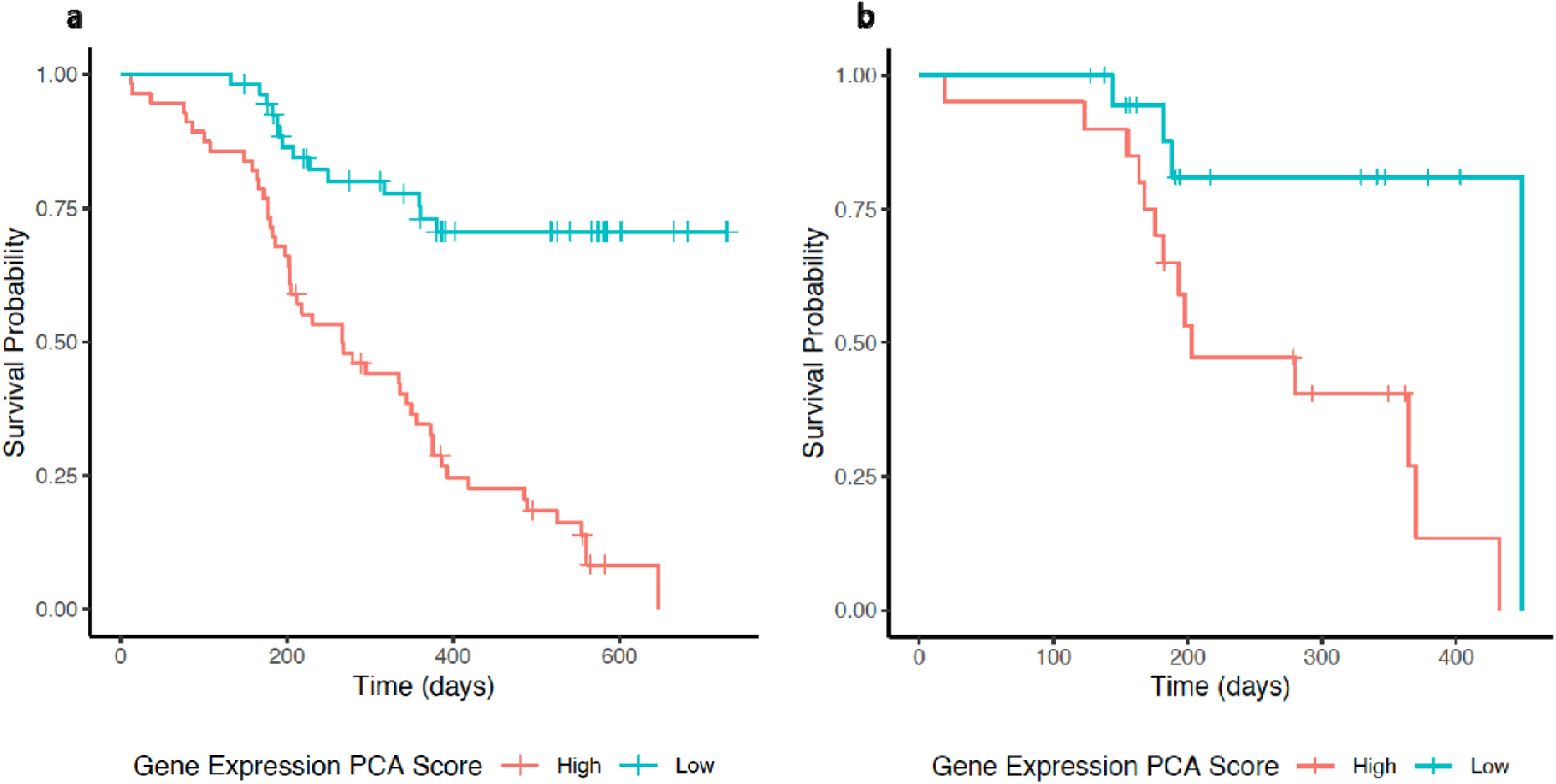
Kaplan-Meier curves displaying high and low gene expression risk scores: (**a**) At baseline, patients with higher gene PCA scores had lower median survival than those with lower scores (267 days vs. not available; *P* = 0.008). (**b**) when assessing the change in gene expression from baseline to 12 months, patients with higher gene PCA scores had lower median survival than those with lower scores (203 days vs. 449 days; *P* = <0.001). PCA = principal component analysis.

## DISCUSSION

In this multicenter study of patients with fHP, we demonstrate that (1) including baseline gene expression signature data leads to a markedly increase in the prediction accuracy of 24-month disease progression compared to using clinical parameters alone, (2) adding the change in gene expression over 12 months to baseline clinical data and changes in FVC% and DTA fibrosis score over that same period, also substantially improve the accuracy of predicting disease progression in the subsequent 12-months, and that (3) a gene PCA score identified fHP patients at greater risk for poor outcomes.

Previous HP transcriptomic studies have been limited by small size, minimal clinical phenotyping, or specimen collection biases, only including subjects with surgical lung biopsy specimens, which are not representative of the broader at-risk HP population or a practical biomarker measurable repeatedly over time. Although molecular phenotyping through gene expression profiling holds promise as a valuable predictor of fHP disease progression, most studies have primarily focused on cross-sectional evaluations to distinguish HP from healthy controls or idiopathic pulmonary fibrosis (IPF). To our knowledge, this is the first prospective transcriptomic study of fHP assessing baseline gene expression and its changes over time to predict disease progression before it occurs.

Although 20 baseline genes were strongly associated with predicting progression in fHP, their biological roles in fHP remain largely unexplored. One exception is glutathione S-transferase Mu 1 (GSTM1), which is known to be linked to susceptibility to pulmonary fibrosis. Variants in GSTM1 have been implicated in pathways related to host defense and oxidative stress that are relevant to the development of pulmonary fibrosis.^19, 20^ However, it still needs to be demonstrated whether these genetic variants influence susceptibility or progression in fHP as well.

The change in the expression profile of the three genes from baseline to 12 months, as identified in our predictive model, associated with subsequent 12-month progression, are novel and has not been previously described in fHP. However, the FAM13A gene is a well-established genetic risk factor for IPF. The single-nucleotide polymorphism (rs2609255) in FAM13A is associated to increased susceptibility to IPF.^21, 22^ While its exact role is still being studied, FAM13A is involved in the Wnt/β-catenin signaling pathway, which is crucial for lung development and is often disrupted in fibrosis. Since IPF and fHP share similar fibrosis and repair pathways, FAM13A’s role may also affect susceptibility to or progression of fHP. SLFN11 is a schlafen family protein regulated by interferons, particularly IFN-γ.^23, 24^ It is expressed in immune cells and may help regulate the cell cycle and activate T-lymphocytes in response to antigens during inflammatory and immune responses. DTWD2 (DTW Domain Containing 2) is an enzyme involved in the post-translational modification of tRNA, specifically in wybutosine biosynthesis, which is key for accurate protein translation.^25^ Disruption of this process may cause cellular stress and may contribute to tissue injury.

Our analysis yielded a gene PCA score that may be helpful for stratifying patients with fHP into low-risk and high-risk groups. The clinical implications of improving outcome prediction and prognostic counseling in HP are substantial. If validated in independent cohorts, this low-risk peripheral blood biomarker score may improve outcome prediction regardless of initial disease presentation, potentially leading to early personalized management strategies.

The strengths of this study include its prospective and multicenter cohort design with a protocol-driven multidisciplinary HP diagnosis to ensure consistency across sites. The study’s aim of assessing peripheral blood gene expression both at baseline and over time was pre-specified before data collection,^8^ thereby minimizing bias in hypothesis testing after the results were known.

However, the study is limited by not been replicated in another independent cohort. While no existing fHP cohort integrates peripheral blood transcriptomics with either disease severity through computer-based methods (radiomics) and longitudinal follow-up, an important next step is to externally test the study findings in an independent population using an alternative NGS platform, focusing on the specific narrow list of transcripts associated with disease progression.

## Conclusions

In a multicenter study of fHP, we demonstrated that a risk-indicative gene expression signature in peripheral blood at baseline, along with its changes over 12 months, predicts disease progression at 24 months and during the subsequent 12 months, respectively. Our analysis suggests that incorporating gene expression data alongside clinical factors significantly outperforms models that rely solely on clinical factors when forecasting disease progression in fHP. Also, this study underscores the potential of molecular phenotyping through gene expression as a promising and valuable point-of-care prognostic assay for the early identification of high-risk patients.

## Data Availability

All data produced in the present study are available upon reasonable request to the authors

## Author’s contributions

Study conception and design: E.R.F.P. Data collection: E.R.F.P., T.M., S.C., T.N.A., A.A., M.B.S. and N.S. Computed tomography analysis: S.M.H. Data analysis: E.R.F.P. S.M.L. and B.V. Initial manuscript draft: E.R.F.P. Manuscript revision for critically important intellectual content: all authors.

